# The study of research barriers from the viewpoint of students and faculty members of the faculty of medicine of Azad University of Najaf Abad

**DOI:** 10.1101/2022.11.12.22282114

**Authors:** Seyed Amirhossein Dormiani Tabatabaei

## Abstract

**Background:** Universities are the most important place for science production and the organization of academic research is one of the most important strategies for community development. The purpose of this study was to identify the barriers of intra-university research in terms of faculty members and students of the faculty of medicine of Islamic Azad University, Najaf Abad.

**Methods & Materials:** This cross-sectional study was carried out in the academic year of 2017-2018 and using a researcher-made questionnaire which included 26 questions in the research barriers section. The questions were divided into five areas: the inability of students to implement the research plan, the importance of not considering the system, the lack of access to facilities and facilities, the lack of cooperation of research centers and the lack of access to research and statistical consultants. Validity of the questionnaire was evaluated by experts and authorities, and reliability was calculated by Cronbach’s alpha of 92.8%. The questionnaire was completed by 49 faculty members and 291 students from the faculty of medicine of Islamic Azad University, Najaf Abad. ANOVA, t-test and Spearman test were used to analyze the data.

**Results:** In this study, the average grades of students in all areas except the lack of access to facilities and facilities more faculty members. The results of this study showed that there is a significant relationship between the two groups’ views on research barriers in all areas except for the lack of cooperation between research centers (P = 0.395).

**Conclusion:** Based on the results of this study, from the viewpoint of faculty and students, research activities are affected by several barriers. The empowerment of human resources, the creation of facilities and facilities, the design of the information and communication system can also be effective in solving the problems and barriers of research.

## Background

The effective role of research in the progress of affairs in recent decades has made research to be considered as an important and key factor in development. Undoubtedly, the effectiveness and fruitfulness of research depends on the level of response to the needs and expectations of the society. Otherwise, research will lose its dynamic and decisive position and become a luxury and unnecessary category, and therefore, the set of research capabilities of the country as a research sector should provide the basic expectations and needs of the society (1). It is certain that in many scientific fields, more than four decades have passed since the beginning of research, and the least expectation is that the product of research is put to the service of implementation and then education and promotion (2). After the training of human resources, research is considered one of the pillars of healthy cultural, social and economic development. In today’s world, only societies can adapt to the rapid developments of science and achieve progress in this way, in which research Institutionalized societies and the main focus of education.

Students toThe active force and researchers of tomorrow can play an important role in the progress of the country (3). Scientific researches, training and clinical treatment are the main and inseparable duties of faculty members (professors) of universities of medical sciences. Meanwhile, students are one of the main pillars and the executive arm and driving engine of research in universities, and professors play pivotal role in encouraging students to conduct student research(4).

Today, countries are classified based on their ability to produce and apply knowledge (5). Undoubtedly, the ability, development and real independence of countries is directly related to the ability to produce science and national development. The production of science and scientific development are known as the driving engine of comprehensive and sustainable development of countries. Undoubtedly, the industrial, economic and social development and progress of any society is dependent on continuous research in all fields (6). In today’s world of knowledge and information advancement, for the advancement of fields that directly or indirectly contribute to patient care, like other academic fields, research is needed, and the expansion of professional knowledge is necessary for continuous improvement in patient care(7). After the training of human resources, research is considered one of the pillars of healthy cultural, social and economic development. In today’s world, only societies can adapt to the rapid developments of science and achieve progress in this way, in which research Institutionalized societies and the main focus of education. Students as an active force and researchers of tomorrow can play an important role in the progress of the country (2). Research in universities of medical sciences is of special importance due to its importance in the field of identifying educational, research, and health problems and issues and providing solutions to eliminate problems related to the health of the society. Research in universities causes Universities are distinguished from other educational centers such as secondary and elementary schools in the educational system (8). Without familiarizing students with research in life and daily learning activities, it will be impossible to cultivate their creative talents in the production of science and participation in the development of the country. In this context, professors play a motivating role in students’ research activities, and by improving their scientific ability, the spirit of research and research of students is strengthened (3).

The results of the studies conducted in Iran have shown that the lack of necessary communication between the university and the research centers, the application of personal tastes in the evaluation of projects and articles, the lack of a specific list of research priorities, the lack of internal information banks related to the conducted research or Currently being implemented, the lack of equipment and facilities needed for research and the difficulty of obtaining them (11, 11) have been obstacles to conducting student research.

. Although the importance and place of research as well as the current inadequacy are approved and agreed by most authorities, but the nature and quality of obstacles and bottlenecks that directly and indirectly affect research activities are not known correctly (12). Recognizing inadequacies is one of the necessary and basic tools that should be available to decision makers and planners in order to make the necessary decisions to achieve goals and improve methods and increase efficiency (13).

growth of students’ participation in research activities is based on strengthening their perspective on the role and place of student research. Officials should establish a reliable and attractive bridge between the classroom and the executive system with accurate and correct planning and provide learning environments and lead students to research with careful supervision (12). In carrying out research activities, there are problems and obstacles that can be solved, and sometimes there are obstacles that block the way of progress. The first step to organize research in the society is to find out the strengths and weaknesses of research programs and recognize the shortcomings. Knowing the research obstacles can facilitate the problem solving process by improving the communication between the researchers and the users of the research results and actually make use of the research findings (14).

Research, as one of the most powerful tools for cultivating potential talents, is one of the most important issues that university planners should pay attention to. Student research causes the student to re-read the relevant texts with special care and to apply various guidelines during practice. to meet the country’s need for researchers in the coming years (21). The development of scientific research and training and experienced experts and competent and prominent experts in every field and creating scientific capability for research and research and the application of theoretical and experimental methods in the field of science and technology will slowly make every country capable of being an importer. The scientific thought and achievements of knowledge and technology of others will become unnecessary (21). On the other hand, it can be acknowledged that the research system is faced with numerous and diverse set of serious issues and problems, which in any case spends an important part of the energy to create, exchange and remove obstacles and correct the functions of the mentioned problems. Finally, it causes a kind of disorder and relative achievement and slows down growth and development (22). Looking at the state of research in the country’s medical science system and comparing it with the state of research in other countries, many disorders in development and research can be seen. Another reason for the need to act on the basis of research is the increase in health care costs; Because conducting research has eliminated some care measures that do not have a favorable outcome and reduces treatment costs. According to Grander, through research and increasing the power and authority of nurses, their job satisfaction increases, which plays an important role in keeping nurses in their profession (7). In any profession, job satisfaction improves the quality of service, and its lack or low level directly or indirectly affects the service. Various studies in Iran show that the level of job satisfaction of nurses and doctors is low, which causes the quality of care provided by the staff to be not optimal, patients recover later and hospital beds are occupied more, and as a result, the cost of the patient and the hospital increase (23 and 24).

One of the effective measures in increasing the job satisfaction of healthcare workers is conducting research by clinical workers, which also reduces costs (15). Systematizing academic research is one of the most important factors affecting university research; Because spreading the spirit of research and creating motivation and desire among academic staff members and researchers towards research depends on creating a system and systematizing research matters and preparing work tools.

The human resources working in the research sector are one of the most important resources of any country for growth and development, and it will not be possible to examine the issue of research without a detailed evaluation of this factor (16). The first step to organize the research in the society is to gain a correct understanding of the available capabilities and possibilities, as well as to understand the weaknesses and strengths of the research programs.

Recognizing the inadequacies and knowing how and how much the goals of the research programs are realized is one of the basic and necessary tools for the decision makers, planners and policy makers of the research, so that through it, the necessary decisions to achieve Objectives, improving methods and increasing efficiency should be adopted. In carrying out research activity, there are problems and obstacles that can be solved, and there are obstacles to progress.

Recognizing research obstacles can facilitate the process of problem solving by improving the communication between researchers and users of research results and actually make use of research findings (17). Factors such as lack of skills in proposal formulation, research implementation, data analysis and interpretation, and essay writing have been reported as individual obstacles to conducting research. The limitation of typing, printer, xerox, research consulting, internet and library are also among the most important factors of people’s dissatisfaction with university research assistance services (15).. It seems necessary to be aware of research obstacles and to overcome them in order to improve the quality and quantity of research. Considering that in developed countries, research has moved from academic centers to clinical centers (12). While in Iran, most of the research is still done by academic staff members.

Considering the importance of student research and the role of professors in guiding these researches and developing students’ talents, as well as the need for planning officials to be aware of the level of professors’ views on student research, this descriptive study aims to evaluate the views of professors and students of the Faculty of Medicine of the Islamic Azad University, Najaf Abad branch. to the obstacles of research and research.

## Methods and Materials

This study is descriptive and cross-sectional on 231 students and 51 faculty members in pre-clinical, health and clinical departments, faculty deans, university presidents, educational, research and financial assistants at the faculty and university level, hospitals directorates Educational and social security clinics and the secretary of the scientific association of university students were conducted in the fall and winter of 2019.

The tool for collecting information in this study is a researcher-made questionnaire, which after reviewing the texts, reviewing the studies and applying the opinions of the professors; Research obstacles in 5 areas were determined as follows:

1. Obstacles related to the lack of ability and sufficient capability of students in the field of research
2. Obstacles related to the lack of access to sufficient facilities and facilities for student research
3. Obstacles related to the lack of proper access to scientific advisors in the field of the implementation process of student research projects in the university.
4. The system does not attach importance to research projects and student theses
5. Obstacles related to the lack of proper cooperation between research centers and hospitals

. Finally, Cronbach’s alpha test was used to determine the validity of the questionnaire and a coefficient of 31.1 was obtained And its reliability was estimated as 1.87 by polling 7 professors and 15 students.

The questionnaire consists of two parts, the first part includes personal and demographic characteristics and the second part includes the questions related to the views of faculty members and the views of students in relation to research obstacles, which are organized in the form of 26 questions. For each question, a score from 0 to 4 (according to the Likert model, from completely disagree to completely agree) was considered. Then, the scores of the questions of each area are added together to get the raw score of that area came and using the following relationship, the score of each area was converted from 0 to 111:

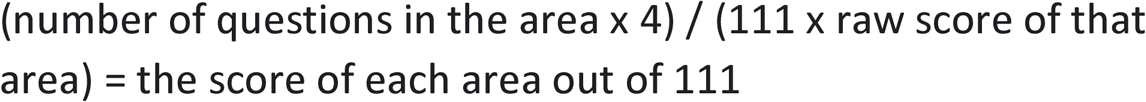

SPSS22 program and independent t-test, Pearson correlation and analysis of variance were used to analyze the data.

## Results

In this study, 67% (34 people) of the studied faculty members were women and the rest were men, and about 77% (255 people) of the students were women and the rest were men. The average age of female faculty members was 46.7±5.3 years, and male faculty members were 43.8±9.2 years old, female students were 5.6±24.6 years old, and male students were 6.6±21 years old.

According to this report, 67.4% of the faculty members had a professional doctorate or higher, 23.3% had a Phd and the rest had a master’s degree.

The average educational experience of women in the university was 16.3±6.5 and that of men was 15.3±7.3.

On average, women have taught 16.5 hours a week and men 15 hours a week during the last year. The average scores of students in all areas except the area of lack of access to facilities and facilities are higher than faculty members, which indicates that students have declared research obstacles more than faculty members. Also, the one-way analysis of variance test shows that there is a significant relationship between the views of the two studied groups regarding research obstacles in all areas, except for the area of non-cooperation of research centers, p=0.04. The independent t test shows that there is a significant difference between the two groups of boys and girls in the area of lack of ability to implement the research plan (P = 118.1). 7.26% of the studied students were married, but the independent t-test shows that between the views of the students Regarding research obstacles, there is no significant relationship with their marital status (P≥0.005). Also, the students who live in the dormitory have a higher average in terms of lack of access to research and statistical consultants and lack of access to facilities and facilities than non-dormitory students, but the test Independent t did not show this difference to be significant (P ≤ 1.15). In this study, students who are working in the fields of the student’s inability to implement the research plan and the lack of cooperation of research centers affiliated to the university have felt the obstacle more (0.05 ≥ p).

This research showed that there is no significant difference between the point of view of students who have had a history of implementing a research project regarding research obstacles, and a group that has not had a history of participation or implementation of a research project (p≥0.05). Also, apart from the area of lack Access to research and statistical advisors. In other cases, the students who are members of the scientific association of the university have declared more barriers to research, but the t-test did not show this difference to be significant (p≥0.05). Also, apart from the lack of access to research and statistical consultants, in the rest of the cases, students with a history of participating in educational workshops related to research have declared the obstacle more, but the independent t test did not show this difference to be significant (p≥0.05) and between There is a significant difference in the opinion score of students who have participated in scientific congresses in the area of non-cooperation of research centers affiliated to the university with the group who have not participated in these assemblies (P=15.1). This research showed that there is a difference between the views of students with a history of publishing scientific articles in terms of lack of access to facilities and facilities compared to students who have no history of publishing articles (P = 112.1).

According to this report, 22 faculty members of the Najaf Abad Faculty of Medicine have independently sent at least one proposal to the research office of the faculty for review and approval, in other words, a total of 30 proposals by 22 faculty members to the research office of the university in the past year. They have sent that about 53% (16 cases) of the proposals have been approved and 10% have been rejected and the rest are under investigation.

About 44.6% of students and 53% of faculty members consider the process of approving proposals to be very long, and 30% of students and 26% of faculty members consider it relatively long.

In other words, 44.6% of students and 53% of faculty members were so dissatisfied with the process of reviewing and approving proposals that they considered it to weaken their motivation in research.

In order, the research obstacles from the faculty members’ point of view were; The student’s lack of ability to implement the research plan, not giving importance to the system, lack of access to facilities and facilities, lack of cooperation of research centers and lack of access to research and statistical consultants. The research obstacles from the students’ point of view were; The student’s lack of ability to implement the research plan, lack of importance to the system, lack of access to facilities and facilities, lack of cooperation of research centers and lack of access to research and statistical consultants.

In this study, the average scores of students in all areas except the area of lack of access to facilities and facilities were higher than faculty members. The results of this study showed that there is a significant relationship between the views of the two studied groups regarding research obstacles in all areas, except the area of non-cooperation of research centers (P = 0.395). 95.9% of faculty members are interested in participating in research projects and 87.8% of them are aware of the benefits of participating in research projects.

83.7% of them have considered professional responsibilities as an obstacle to carry out research projects.

52.6% of students were interested in participating in research projects, while only 22.7% of them knew about the benefits of participating in research projects.

## Conclusion

Scientific researches, training and clinical treatment are the main and inseparable duties of faculty members (professors) of universities of medical sciences. Meanwhile, students are one of the main pillars and the executive arm and driving engine of research in universities, and professors play a pivotal role in encouraging students to conduct student research(3).

Today, countries are classified based on their ability to produce and apply knowledge (4). Undoubtedly, the ability, development and real independence of countries is directly related to the ability to produce science and national development. The production of science and scientific development are known as the driving engine of comprehensive and sustainable development of countries. Undoubtedly, the industrial, economic and social development and progress of any society is dependent on continuous research in all fields (5).

The growth of students’ participation in research activities is based on strengthening their views on the role and place of student research. The officials should establish a reliable and attractive bridge between the classroom and the executive body with accurate and correct planning and provide learning environments. And with careful supervision, lead students to research (6). In carrying out research activities, there are problems and obstacles that can be solved, and sometimes there are obstacles that block the way of progress. The first step to organize research in society is to find out the strengths and weaknesses of research programs and recognize their inadequacies. Recognizing research obstacles can facilitate the problem solving process by improving the communication between researchers and users of research results and actually lead to the use of research findings (17).. Knowing the research obstacles can facilitate the problem solving process by improving the communication between the researchers and the users of the research results, and actually lead to the use of the research findings, and not paying attention to this matter causes interruptions in conducting research, and perhaps these interruptions cause losses. They cause irreparable damage that makes the research unsuccessful (8). The growth of students’ participation in research activities is based on strengthening their perspective on the role and place of student research. The authorities should establish a reliable and attractive bridge between the classroom and the executive body with accurate and correct planning and provide learning environments and lead students to research with careful supervision (6). In carrying out research activities, there are problems and obstacles that can be solved, and sometimes there are obstacles to progress. The first step to organize research in the society is to find out the weaknesses and strengths of research programs and recognize the shortcomings. Knowing the obstacles to research can be improved The communication between the researchers and the users of the research results will facilitate the problem solving process and actually lead to the use of the research findings (7). The results of the studies conducted in Iran have shown that the lack of necessary communication between the university and the research centers, the application of personal tastes in the evaluation of projects and articles, the lack of a specific list of research priorities, the lack of internal information banks related to the conducted research or Currently, the lack of equipment and facilities needed for research and the difficulty of obtaining them have been obstacles to conducting student research(4). Although the importance and position of research as well as the current inadequacy are emphasized and agreed by most officials.

The nature and quality of obstacles and bottlenecks that directly and indirectly affect research activities are not known correctly. Recognizing inadequacies is one of the necessary and basic tools that should be available to decision makers and planners so that necessary decisions can be made to achieve goals and improve methods and increase efficiency (4). This study aims to determine research obstacles from the point of view of students and faculty members of the Faculty of Medicine,Najaf Abad Azad University. In this study, the barriers to research from the perspective of faculty members were: Lack of student’s ability to implement the research project (15.37±16.6), not giving importance to the system (7.2±75.16), lack of access to facilities and facilities (52.15±3.2), non-cooperation of centers research (3.2±57.14) and lack of access to research and statistical consultants (23.11±25.2). The research obstacles from the students’ point of view were; Lack of student’s ability to implement the research project (71.37±32.4), not giving importance to the system (13.18±2.8), lack of access to facilities and facilities (23.15±7.2), non-cooperation of centers research (15.2±1.15) and lack of access to research and statistical consultants (4.11±3.2).

In this study, the average scores of students in all areas except the area of lack of access to facilities and facilities were higher than that of faculty members. The results of this study showed that there is a significant relationship between the views of the two studied groups regarding research obstacles in all areas, except the area of non-cooperation of research centers (P = 0.395). In a research conducted by Nik Afrooz et al., the unavailability of consulting forces, the weakness of consulting forces, the lack of facilities and equipment, and the lack of motivation on the part of officials and professors are the most common obstacles in the field of organizational obstacles. (35).In Abedini’s study, the obstacles were as follows: Lack of necessary facilities and equipment in 22 cases (8.32 percent), unavailability of consulting forces in 21 cases (23.3 percent) (3). In the study of Alamdari in Yasouj, the lack of necessary facilities and equipment was mentioned as the biggest obstacle to research by the organization (7). In the study of nature, lack of motivation in researchers, lack of time and busyness, and cumbersome administrative regulations have been among the most important obstacles to conducting research.

Seresshti stated in his article that the need to remove the obstacles is felt by the vice president of research of the university, the officials of hospitals and healthcare centers. The results of this part of the present study are consistent with similar studies. The findings of this study show that the average score of male students was lower than that of female students in the area of lack of ability to implement the research plan. In other words, female students in these areas have declared more obstacles (P = 118.1). The results of a study conducted on research barriers in Isfahan University showed that there is a significant difference between the score of personal barriers and organizational barriers in both sexes. This means that opposition to organizational obstacles is much more than opposition to personal obstacles. The results of the present study are consistent with the study of Kianpour et al. only in the section of students’ views regarding the inability to implement a research plan, and in other areas, no significant difference was observed in the present study. This difference between the present study and the study of Kianpur can be caused by the difference in the capacities and research possibilities of the two universities.

The results of Sabzevari’s study on medical students in Kerman showed that the average score of individual barriers to conducting research is higher than organizational barriers (6); which is different from the results of the present study, which can be caused by the difference in the research population of the two studies. The present study was conducted only on medical students and Sabzevari’s study was conducted on medical students. It can also be due to different facilities in two universities.

In a research conducted by Sarshti et al. at Shahrekord University of Medical Sciences, the highest average score of obstacles was related to lack of time and lack of sufficient motivation on the part of the authorities, lack of advisory staff and lack of facilities, which is different from the present study in terms of ranking. Is. This difference can be due to the different facilities and research capacities of the two universities (6). Also, in Sereshti’s study, the students who are working in the fields of the student’s inability to implement the research plan and the lack of cooperation of the university-affiliated research centers have felt more obstacles, which is consistent with the present study. The results of this study showed that there is no significant difference between the point of view of students who have had a history of implementing a research project regarding research obstacles, and a group that has not had a history of participating or implementing a research project. In his study, Roxburgh identifies lack of time, lack of support system, and insufficient knowledge and skills in the field of research methodology as the most important obstacles to conducting research. In the Myles study, there is a difference between the views of students who have had a history of conducting research and a group who have not had a history of conducting research and research. This difference between the results of the present study and the Roxburgh study can be caused by the difference in the research capacities of the two studies (14). The independent t-test in this study showed that there is no relationship between students’ views on research obstacles and their marital status (P≥0.005). This part of the results of the study is consistent with the results of Sareshti’s study at Shahrekord University of Medical Sciences and the results of Leavid and Baikal’s study.

The results of this study show that students who live in the dormitory have a higher average in terms of lack of access to research and statistical consultants and lack of access to facilities and facilities than non-dormitory students, but the independent t-test did not show this difference to be significant (1/ 15 ≤ P). This indicates that the students living in the dormitory have more access to research experts and statistical consultants because they spend most of their time in the university, university sites and the library. which is consistent with the results of Javadian’s studies (11).

The present study showed that the students who are working, in the areas of lack of ability of the student to carry out the research project and the lack of cooperation of the research centers affiliated to the university have felt the obstacle more (p≥0.05). This part of the study is consistent with the results of Sarshti et al.’s study (15).

The results of this study regarding the history of research project implementation and its relationship with research obstacles indicate that the difference between the score of students who have had a history of research project implementation and research obstacles, and a group that has not had a history of participation or research project implementation. There is no significant difference (p≥0.05). This part of the study with the results of Sabzevari’s study which stated; People who did not have a research project had more individual and organizational obstacles average score, which is inconsistent and can be caused by the difference in the size of the samples and the difference in the existing organizational policies in the two universities.

The results of our study showed that there is a significant difference between the point of view of students who have participated in scientific congresses in the area of non-cooperation of university-affiliated research centers and the group who have not participated in these assemblies (P = 15.15).). This part of the study is also based on the results of Sabzevari’s study which stated; People who did not have a research plan had higher average scores of individual and organizational obstacles, it is consistent (26).

Regarding the announcement of research obstacles in a group of students who have passed the training workshop and comparing it with the group that did not pass these workshops; The results of this study indicate that, apart from the area of lack of access to research and statistical consultants, in other cases, students with a history of participating in educational workshops related to research have declared the obstacle more. This difference can indicate that the group who passed the workshops have a tendency to cooperate with their professors in the implementation of the research, therefore, during the implementation of their research project, the obstacles i.e. lack of access to the facilities and facilities needed for their project and attach importance They will face the failure of the system and feel the obstacles in these two areas more than the other group. This part of the results of the present study is consistent with the results of Sarshti and Roxburgh studies.

The results of this study indicate the existence of organizational barriers and personal barriers to conducting research in Najaf Abad Azad University from the point of view of medical students and faculty members, which can be corrected and controlled by professors, faculty members, and university officials. Creating motivation on the part of officials and professors can reduce the obstacles of student research to some extent. It seems that teaching in an applied manner, in such a way that the student is really involved in the research process and creating the necessary platforms in this regard, is effective.

Also, the results of this research show that most professors of the Faculty of Medicine of Najaf Abad Azad University are interested in doing research work, but for several reasons, including the lack of familiarity and experience of students with the principles of research and the lack of sufficient facilities, they are not very willing to guide student research.

According to the results of this research, it is suggested that;

1. Necessary planning should be done to hold educational workshops related to research. 2. Necessary measures to increase the cooperation of faculty members and graduate students with Medical students to familiarize students with the research process and the implementation of research projects.
3. Necessary planning should be done in order to familiarize research students with the principles of research and its implementation from the level of basic sciences.
4. Necessary planning should be done to form research cores in universities and hospitals, as well as to strengthen team research.
5. Studies with the same objective as the present study should be conducted at the university level and survey all fields and departments.

## Data Availability

All data produced in the present study are available upon reasonable request to the authors.

https://ethics.research.ac.ir/EthicsProposalView.php?id=74441

## References

1. Marusic B. Academic medicine: one job or three? CMJ 2014; 45(3): 243–244.

2. Salem safi R, Ashrafrezaei N, Moshiri Z, Shaykhi N, Baniadam Study of views of faculty members about research barriers in Urmea university of medical sciences. Journal of faculty Nursing and Midwifery of Urmea University. 2017;7(3):142–150.

3. Fealy S, Pezeshkirad Gh, Chizari M. Study of the factors affecting students participate in research and knowledge translation. Quarterly of research and programming in education. 2016;42.

4. Alaei M, Azami A. Study of student attitudes toward research in Eilam university of medical sciences. Journal of Eilam university of medical sciences. 2014;12(42-43):39–42.

5. Arab mokhtari R, Naderian Jahromy M, Soltan Hosieny M. Identify barriers and problems in Research in exercise disciplinary in the country’s public universities (Dissertation). Faculty of Physical Education and Sport Sciences. 2017.

6. kianpour M, Bahmanziari P, Atri S. Survey of research problems from the viewpoints of managers. Scientific staffs and research experts in Isfahan university of medical sciences. Iranian journal of nursing and midwifery research. 2016;(28):23–24.

7. Farmanbar R, Askari F. Study of barriers related to research: views of faculty members in Gilan university of medical sciences. Journal of Medical faculty of Gilan university of medical sciences. 2015;14(54):84–91.

8. Lioyd T, Philips BR, Aber RC. Factors that influence doctors’participation in clinical research. Med Educ.2014;38(8):848–51.

9. Bickel J. Women in academic medicine. J Am Med Womens Assoc. 2014;55(1)10–2.

10. Couvillon JS. How to promote or implement evidenced-based practice in clinical setting. Home Health Care Manag Prac. 2015;17(4):269–72.

11. Roxburgh M. An exploration of factors which constrain nurses fron research participation. J Clin Nurs. 2016;15(5)535–45.

12. Young Ra, Dehaven MJ, Passmore C, Baumer JG. Research participation, protected time and research output by family physicians in family medicie residencies. Fam Med. 2016;38)5(:8–341.

13. Aronstam RS, Hoey K, Frieije JE. Physician participation in clincal research at Guthrie health. Guthrie J. 2014;721(3):66–71.

14. Beqaei A., Owais Qaran S., Noot Dost B., Mohammadi M., Asgari Moghadam M., Shekarchizadeh A. Obstacles to research in medical sciences from the point of view of research officials and faculty members: collection of articles of the first scientific conference on research issues of the country, May 28 and 23, 1332; 154–143.

15. Research on research problems in Iran: Journal of Research and Industry. 1334; 7 (15): 3–7.

16. Panjeh Shahin MR, Mola A, Nabavi Zadeh A, Tavakoli AR. Effective factors on research activities of student researchers of Shiraz University. Proceedings of Cardiovascular Congress of Shiraz University of Medical Sciences 2016; Page 94.

17. Nikafrooz Laila. Examining the factors preventing the implementation of research projects from the perspective of students. Journal of Fasa University of Medical Sciences, No. 6, Summer 1333, 313-313.33

18. Sereshti manijeh. Barriers to conducting research from the point of view of professors and staff of Shahrekord University of Medical Sciences. Journal of Education Strategies. Third period, number 2, summer 1336: 51–57.

19. Aronstam RS, Hoey K, Frieije JE. Physician participation in clincal research at Guthrie health. Guthrie J. 2015;721(3):66–71.

20. Rudd FG, Harasym PH, Mandin H, Lorscheider FL. Development and evaluation of a research project

21. Smith KJ, Mohn K, Pinevich AJ, Nasca TJ. Residency requirements for scholarly activity. Acad Med 2016;71(3): 214.

22. Haynes B, Haines A. Barriers & Bridge to evidence based clinicalpractice. British Medical Journal 2012;273–374.

23. Hamilton, GA: Two faces of nurse faculty: teacher &research, jurnal of Advanced nursing, 2016, 11(8):217–223.

24. Soltani A. Examining the current situation in the future perspective of research. Investigating the cultural barriers of research in the country: Proceedings of the first scientific conference on research issues in the country, 28 and 23 May 1332: 311–311.

25. Yaqoubi, Tahereh. Examining the obstacles and problems of implementing research projects from the point of view of faculty members Mazandaran University of Medical Sciences. Journal of the University of Medical Sciences (special issue of the fourth national conference Medical Education), 1333 p. 186.

26. Salem safi R, Ashrafrezaei N, Moshiri Z, Shaykhi N, Baniadam Study of views of faculty members about research barriers in Urmea university of medical sciences. Journal of faculty Nursing and Midwifery of Urmea University. 2011;7(3):142–150.

27. Haynes B, Haines A. Barriers & Bridge to evidence based clinicalpractice. British Medical Journal 2017;273–374.

28. Hamilton, GA: Two faces of nurse faculty: teacher &research, jurnal of Advanced nursing, 2016,11(8):217–223.

29. Marusic B. Academic medicine: one job or three? CMJ 2014; 45(3): 243–244.

30. Asifzadeh Saeed, Piri Zakieh. publication; Key activity in knowledge management. Journal of Babol University of Medical Sciences. 1332; 8(4): 54–62.

31. Mousavi Mohadi Ali Akbar. Production of science and its criteria. the approach 1331; 28: 144–142.

32. Kianpour M, Bahmanziari P, Atri S. Survey of research problems from the viewpoints of managers. Scientific staffs and research experts in Isfahan university of medical sciences. Iranian journal of nursing and midwifery research. 2016;(28):23–24.

33. Alamdari A, Afshoon A. Barriers to research activities: Views of faculty members in Yasuoj university of medical sciences. Journal of Armaghane danesh. 2012;29:27–34.

34. Halid D, Darrell RI. Determinant of Research Productivity in Higher Education. Research in Higher Education 2016, 36(6):607–31.

35. Abedini S, Abedini S, Kamalzadeh H, Momeni A, Zare Sh. Views of faculty members in Yasouj university of medical sciences about the barriers to research activities. Dena Quarterly. 2017; 4(1 & 2):51–58

